# Cognitive performance, frailty and functional dependence in community-dwelling older adults: Results of the FREEDOM cohort

**DOI:** 10.64898/2026.07.04.26357264

**Authors:** Ludwig Mounsamy, Caroline Gayot, Cécile Laubarie-Mouret, Noëlle Cardinaud, Nathalie Dumoitier, Michel Druet-Cabanac, Alexandre Lagrange, Benjamin Festou, Karen Rudelle, Achille Tchalla

## Abstract

**BACKGROUND:** Aging is associated with a progressive decline in cognitive performance and functional autonomy, both closely related to frailty. Understanding the interrelation between these domains is essential to identify modifiable factors influencing cognitive impairment in older adults.

**OBJECTIVES:** To evaluate the relationship between physical frailty, cognitive performance, and functional dependence, and to identify sociodemographic and clinical variables associated with cognitive impairment in community-dwelling older adults.

**DESIGN:** Cross-sectional study.

**SETTING:** FREEDOM-LNA cohort, a population-based study conducted by the University Hospital of Limoges, France.

**PARTICIPANTS:** A total of 753 community-dwelling older adults aged ≥75 years, or ≥65 years with at least two comorbidities, were included.

**MEASUREMENTS:** Cognitive function was assessed using the Mini Mental State Examination (MMSE), 5-word test (5WT), clock drawing test (CDT), and verbal fluency tests. Frailty was defined according to Fried’s physical criteria, and functional independence was evaluated using ADL and IADL scales. Sociodemographic, clinical, and lifestyle factors were analyzed using multivariate models to identify predictors of cognitive impairment. RESULTS: Of the participants, 34.4% had a pathologic MMSE, 46.0% failed the CDT, 68.0% the verbal fluency test, and 17.0% the 5WT. Cognitive performance was significantly lower among frail compared to prefrail and robust individuals. Older adults with pathologic cognition were more frequently dependent in activities of daily living. Independent predictors of poor cognitive performance included non-modifiable factors (age, sex, education) and modifiable ones (low BMI, hypertension, alcohol consumption, smoking, and polypharmacy). CONCLUSIONS: Cognitive impairment was highly prevalent among frail older adults and was strongly associated with loss of independence. Interventions targeting modifiable risk factors such as low BMI, hypertension, alcohol consumption, and smoking may help preserve cognitive and functional abilities in aging populations.

Interventions to improve BMI and reduce alcohol consumption, smoking, and hypertension may preserve cognition in older adults. (150 words)

## 1. Introduction

With the aging population and increasing life expectancy, a large number of older individuals live with physical disability causing difficulties in daily activities. The maintenance of age-sensitive cognitive functions is a criterion of successful aging [Nyberg, 2019; Friedman, 2019]. Older adults may experience decline in the cognitive domains, attention, memory, executive cognitive function, language production, and visuospatial abilities, affecting basic and instrumental daily activities [Murman, 2015]. Cognitive impairment in older adults is frequently associated with increased risks of adverse health-related outcomes including falls, dementia, hospitalizations, and mortality. Individuals with low cognitive impairment are at higher risk of institutionalization than non-impaired older individuals [Avila-Fuenes, 2009; Houles, 2012]. Evidence from observational studies supports a temporal association between frailty, cognitive impairment, and dementia; epidemiological evidence also suggests that cognitive impairment is a component of frailty [Cleg, 2013; Khelaidity, 2013; Halil, 2015].

Frailty is defined as a clinical syndrome in which a decreased reserve and resistance to internal or external stressors, resulting from cumulative decline across multiple physiological systems, increases vulnerability to adverse health-related outcomes including falls, disability, hospitalization, institutionalization, mortality, and cognitive-related outcomes (dementia and late-life cognitive disorders) [Fried, 2001; Panza, 2018]. Frail individuals with cognitive impairments may be more likely to experience disability and hospitalization compared to those without cognitive impairment [Avila-Funes, 2009].

Frailty may be assessed using physical criteria, including weak muscle strength, slow gait speed, unintentional weight loss, exhaustion, and low physical activity. Some of these physical criteria are independently associated with cognitive function [Robertson, 2014]. Frailty is also associated with decreased cognitive performance over time. Late-life progressive physical frailty and declining cognition may be strongly correlated in part because they share a common pathologic basis [Buchman, 2014; Searle, 2015]. Consequently, cognitive frailty has been defined as the simultaneous presence of physical frailty and mild cognitive impairment in the absence of dementia or other pre-existing brain disorders [Kelaiditi, 2013]. Physical and cognitive frailty are potentially reversible and frailty status may be a modifiable target in early cognitive impairment [Borges, 2019; Negm, 2019; Panza, 2018].

Interventions to preserve cognition, robustness, and loss of independence are necessary. Given the apparent interrelation between physical frailty and cognitive impairment, efforts should focus on understanding this relationship to identify individuals with potentially reversible cognitive impairment [Sargent, 2017]. Interventions should be implemented before neuronal damage becomes irreversible [Oliveira, 2018].

Monitoring frailty indicators, including cognitive function, in older individuals is recommended to identify those who could benefit from disability prevention programs. To provide insight into the development of frailty in the elderly, we implemented the FREEDOM-LNA cohort of community-dwelling older adults to study aging and identify predictive factors of frailty, loss of independence, and cognitive function [Boyer, 2022]. In this cross-sectional analysis, we evaluated the prevalence of pathologic cognitive function assessed using a battery of neuropsychological tests in relation to frailty status and functional capacity in older adults of the FREEDOM-LNA cohort. We also identified sociodemographic, medical, and lifestyle variables predictive of cognitive impairment.

## 2. Materials and méthods

### 2.1 Study design

This study was a cross-sectional analysis of the FREEDOM-LNA cohort [Boyer *et al*., 2022]. Briefly, the FREEDOM-LNA longitudinal observational study is conducted by the UPSAV (*Unité de Prévention de Suivi et d’Analyse du Vieillissement*) at the University Hospital of Limoges, France. Overall, 1085 community-dwelling adults over 75 years of age, or over 65 years of age with at least two comorbidities were analyzed. The characteristics of the FREEDOM cohort were reported elsewhere [Boyer *et al*., 2022]. The study protocol was reviewed and approved by the local institutional review board (CEREES, Limoges; TPS 429669) and by the French data protection authority (CNIL) to ensure protection of individualized data according to French law. Informed consent for data processing was obtained from all participants (or their legal representatives). All procedures were carried out in accordance with the Declaration of Helsinki and its amendments.

### 2.2 Measurements

A comprehensive geriatric assessment was performed at home and questionnaires on sociodemographic characteristics, medical history, nutritional status, and physical status were administered by health professionals (geriatric physicians or trained nurses) [Boyer *et al*., 2022].

#### 2.2.1 Cognition and mood

Neurocognitive domains – verbal memory, immediate memory, and executive functioning – were assessed using neuropsychological tests: the Mini Mental State Examination (MMSE) questionnaire [Folstein, 1975], the 5-word test (5WT) [Dubois, 2002], the clock drawing test (CDT) [Schulman, 2000], the Controlled Word Association Test [Malek-Ahamadi, 2011], and the Category Naming Test [Isaacs, 1973]. Participants were considered to have a cognitive deficit if the MMSE score was ≤ 20 in people with a low educational level, ≤ 23 in people with a medium educational level, and ≤ 26 in people with a high educational level. The 5WT ranges from 0 to 10, and a score < 9 indicates a deficit in memory performance. The CDT was used to assess executive function; it is a simple cognitive screening tool that is easy to administer and has high sensitivity and specificity to detect dementia [Shulman, 2000]. Participants were presented with a pre-drawn circle and asked to place the numerals correctly and draw the clock hands set to a time indicated by the examiner. The test was scored as 0 (failure) or 1 (success).

The 30-item Geriatric Depression Scale (GDS) was used to assess depression. In the GDS, subjects are asked to respond yes or no to questions on how they felt over the past week. Scores ranging from 0 to 5 indicate normal mood; scores of 5 to 9 indicate mild depressive symptoms, and scores of ≥ 10 indicate severe depressive symptoms [Yesavage, 1983].

#### 2.2.2 Measurement of frailty

Frailty was defined as three or more of five phenotypic Fried criteria: weakness (grip strength of the dominant hand < 20%), slowness (walking speed < 20% of normal), low level of physical activity (< 20% of energy expenditure), low energy or self-reported exhaustion, and unintentional weight loss (4 to 5 kg since the previous year) [Fried, 2001]. Subjects were considered frail if at least three criteria were present, pre-frail if one or two criteria, and robust if no criterion was present.

#### 2.2.3 Measurement of functional autonomy

Functional autonomy was assessed using the six-item activities of daily living (ADL) [Katz, 1983], and eight-item instrumental activities of daily living (IADL) tests [Lawton, 1970]. For each item, the disability was scored as 1 (independent), 0.5 (needs help), or 0 (dependent). An ADL score ≤ 5 indicates a subject dependent for daily activities and IADL ≤ 7 indicates dependence for instrumental daily activities.

### 2.3 Covariates

Covariates deemed to influence cognitive and psychosocial functions included sociodemographic variables (age, sex, and educational level [low, intermediate, or high]), cardiovascular risk factors (hypertension, dyslipidemia, and diabetes), polypharmacy (≥ 5 medications per day), lifestyle (smoking, alcohol consumption, and living arrangement), and body mass index (BMI; < 21; 21–25; 25–30 and > 30 kg/m^2^).

### 2.7 Statistical analysis

Statistical analysis was performed using SAS software, version 9.4 (SAS Institute, Cary, NC). Quantitative variables are presented as means, standard deviations (SD), medians, and minimum and maximum values. Qualitative data are presented as numbers and percentages. Missing data were not replaced, and percentages were calculated without accounting for missing data.

Univariate analyses using linear regression models were performed to assess the significance of the associations of covariates with cognitive scores. Multivariate models were implemented to identify independent covariates. We applied a backward stepwise selection controlled for all factors with a P < 0.20 in univariate models to select significant factors (at the 5% level).

The association between pathologic cognition as a binary variable (yes or no) and frailty was described in contingency tables and evaluated by chi-squared test or Fisher exact test (if fewer than five cases in one category). The same statistical methodology was used to evaluate the association between pathologic cognition and the ADL and IADL scales. All tests were bilateral and considered significant at an alpha level of 0.05 (P < 0.05).

## 3. Results

Table 1 lists the baseline characteristics of the 753 subjects (mean age 83.1 ± 5.8 years, 68% females). Of these subjects, 240 (31.9%) were frail, 439 (58.3%) were pre-frail, and 74 (9.8%) were non-frail (robust) according to Fried’s criteria. Limitation in at least one ADL (ADL < 5) was present in 456 (60.6%) subjects and in at least one IADL in 605 (80.3%) subjects.

**Table 1.**
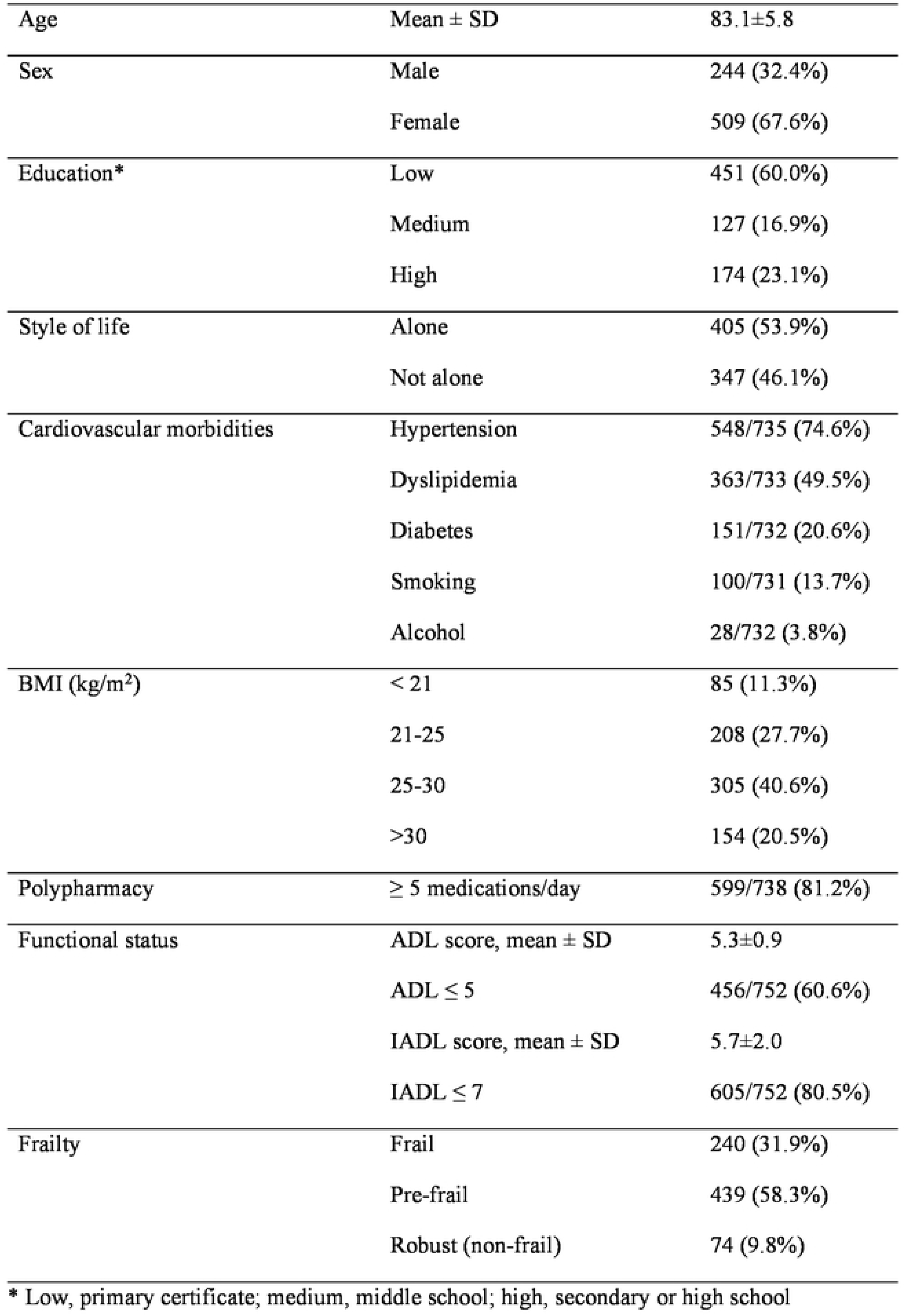
Characteristics of the study population (N = 753 subjects)

|  |  |  |
| --- | --- | --- |
| Age | Mean ± SD | 83.1±5.8 |
| Sex | Male | 244 (32.4%) |
|  | Female | 509 (67.6%) |
| Education* | Low | 451 (60.0%) |
|  | Medium | 127 (16.9%) |
|  | High | 174 (23.1%) |
| Style of life | Alone | 405 (53.9%) |
|  | Not alone | 347 (46.1%) |
| Cardiovascular morbidities | Hypertension | 548/735 (74.6%) |
|  | Dyslipidemia | 363/733 (49.5%) |
|  | Diabetes | 151/732 (20.6%) |
|  | Smoking | 100/731 (13.7%) |
|  | Alcohol | 28/732 (3.8%) |
| BMI (kg/m<sup>2</sup>) | < 21 | 85 (11.3%) |
|  | 21-25 | 208 (27.7%) |
|  | 25-30 | 305 (40.6%) |
|  | >30 | 154 (20.5%) |
| Polypharmacy | $\geq 5$ medications/day | 599/738 (81.2%) |
| Functional status | ADL score, mean ± SD | 5.3±0.9 |
| | ADL $\leq 5$ | 456/752 (60.6%) |
| | IADL score, mean ± SD | 5.7±2.0 |
| | IADL $\leq 7$ | 605/752 (80.5%) |
| Frailty | Frail | 240 (31.9%) |
|  | Pre-frail | 439 (58.3%) |
|  | Robust (non-frail) | 74 (9.8%) |

\* Low, primary certificate; medium, middle school; high, secondary or high school

Cognitive function was assessed using the MMSE, 5WT, CDT, and verbal fluency (Table 2). The MMSE total score was 25.0 ± 4.3, and 34.4% (245/713) of the subjects had a pathologic MMSE after adjustment for education. Additionally, 106 of 623 (17.0%) subjects had a 5WT < 9, indicating poor memory performance; 291 of 632 (46.0%) failed the clock-drawing test, suggesting some executive dysfunction. Based on the verbal fluency tests, 379 of 557 (68.0%) subjects had at least one categorial or literal fluency considered pathologic. The GDS questionnaire was completed by 634 subjects with a mean score of 9.6 ± 5.4; 302 of the 634 (47.6%) subjects had a GDS score ≥ 10, indicating depression.

**Table 2.** Cognitive function and depression.

|  |  |  |
| --- | --- | --- |
| MMSE | N | 713 |
| Total score | Mean ± SD | 25.0±4.3 |
|  | Pathologic MMS* | 245 (34.4%) |
| 5WT | N | 623 |
| | Mean ± SD | 9.2±1.6 |
|  | 5WT < 9 | 106 (17.0%) |
| Clock drawing test | N | 632 |
|  | Failure | 291 (46.0%) |
| Verbal fluency | N | 557 |
|  | Pathologic categorial fluency | 334 (60.0%) |
|  | Pathologic literal fluency | 168 (30.2%) |
|  | At least one pathologic verbal fluency<br>(Suspected cognitive impairment) | 379 (68.0%) |
| GDS | N | 634 |
| | Mean ± SD | 9.6±5.4 |
|  | GDS > 9 | 302 (47.6%) |

\* MMSE < 24–27 according to educational level.

### 3.1 Relationship between frailty and cognition and depression

As shown in Figure 1, cognitive performance differed significantly according to frailty status. Of the frail subjects, 48.2% had a pathologic MMSE *versus* 30.2% (P < 0.001) of pre-frail subjects and 15.7% of frail subjects; 19.7% of frail subjects had a 5WT < 9 *versus* 8.1% of non-frail subjects (P = 0.034) and 17.2% of pre-frail subjects (P = 0.474); 59.7% of frail subjects failed the CTD test *versus* 26.6% of non-frail (robust) subjects (P<0.001) and 42.7% of pre-frail subjects (P < 0.001). Additionally, 77.3% of frail subjects had some pathologic verbal fluency *versus* 47.5% of non-frail subjects (P < 0.001) and 67.2% of pre-frail subjects (P = 0.020). With regard to depression (Figure 2), 70.5% of the frail subjects had a GDS score > 9 *versus* 24.2% (P < 0.001) of non-frail (robust) subjects and 40.2% (P < 0.001) of pre-frail subjects.

**Figure 1.**
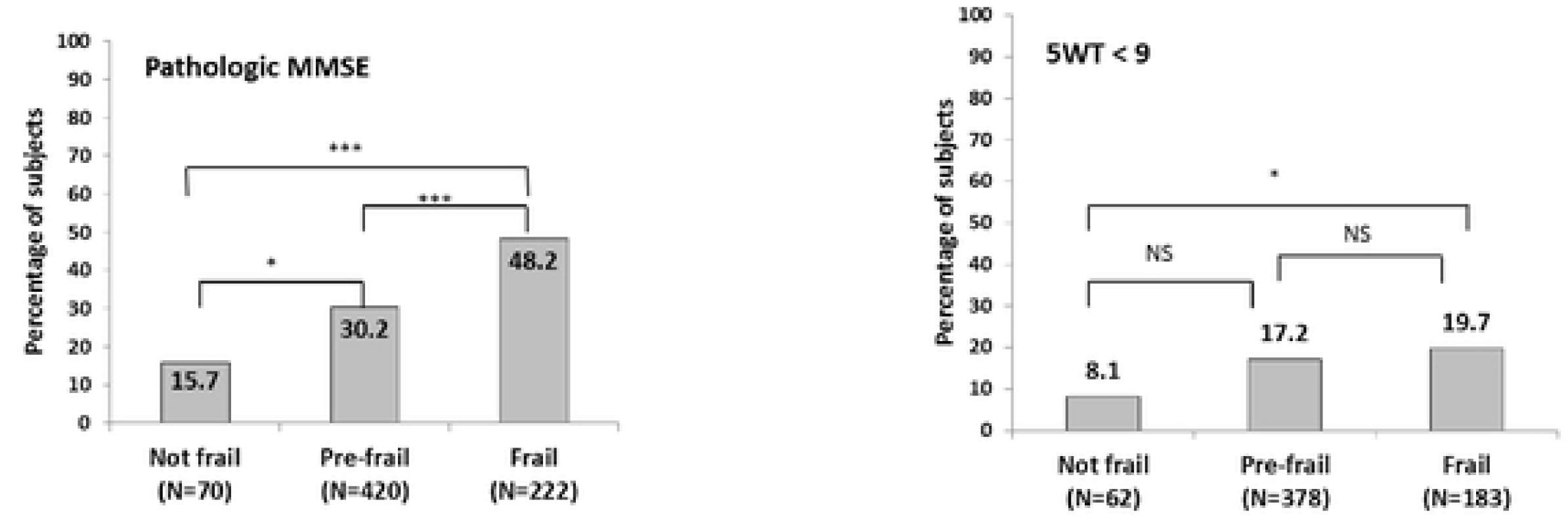
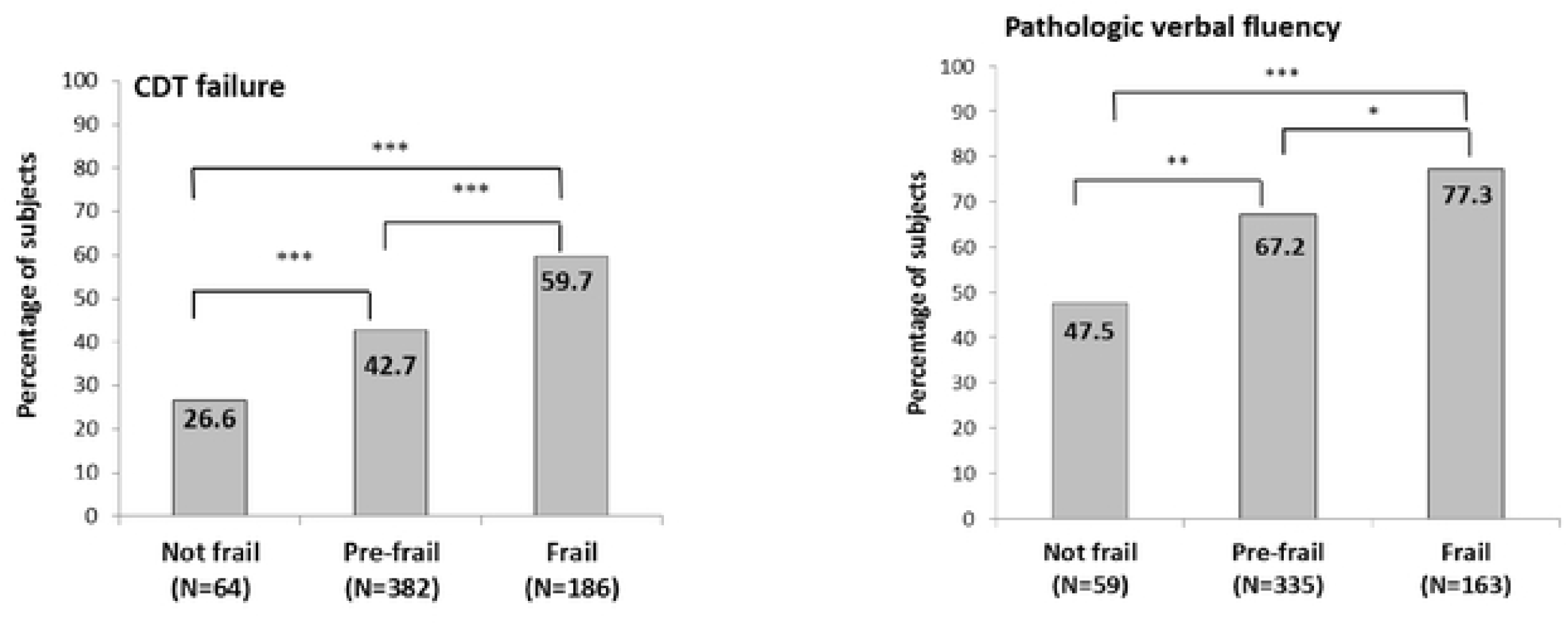
Relationship between frailty and 1>athologic cognition

**Figure 2.**
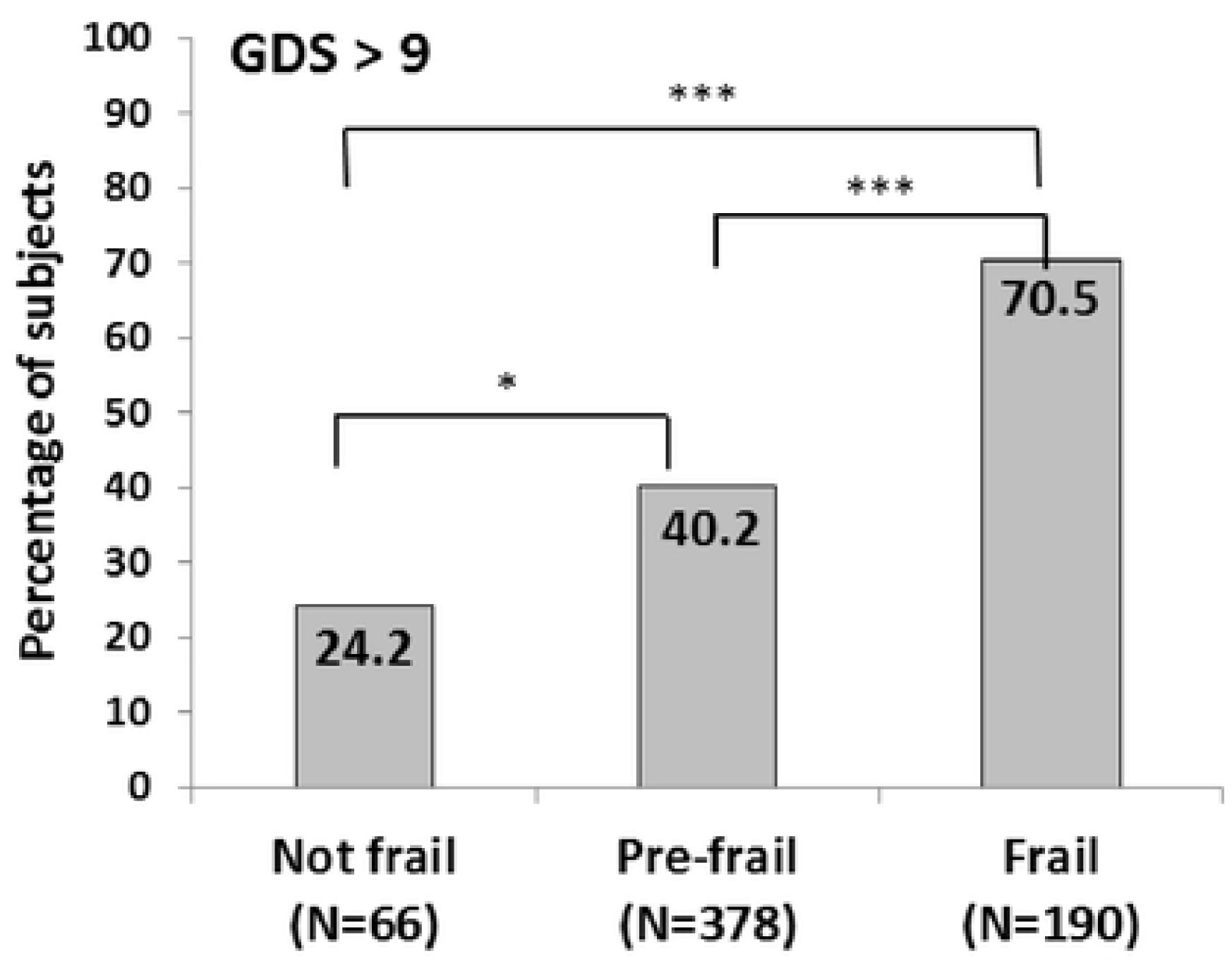
Relationship between frailty and depression

### 3.2 Relationship between pathologic cognition and limitation in activities of daily living

As shown in Figure 3, subjects with pathologic cognition were significantly more frequently affected in ADL and IADL compared to subjects without pathologic cognition. Subjects with depression (GDS score > 9) were significantly more frequently affected in ADL and IADL compared to subjects without depression.

**Figure 3.**
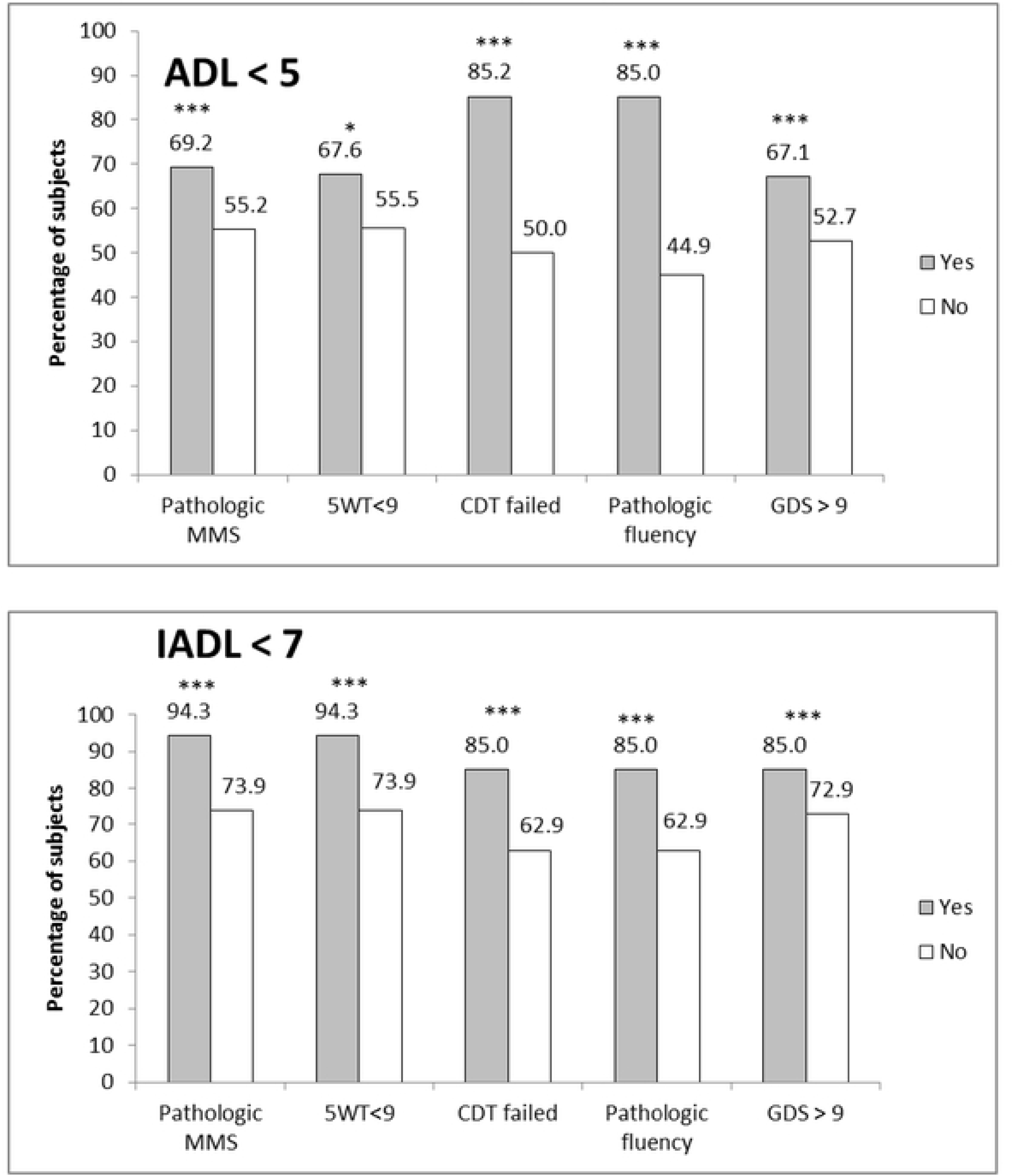
Relationship between pathologic cognition (cognitive deficit) and loss of autonomy (limitations in activities of daily living)

### 3.3 Factors associated with loss of cognition and depression

Multivariate analyses identified significant prognostic factors for pathologic cognition and depression (Table 3). Increasing age was associated with a higher likelihood of pathologic MMSE (adjusted OR = 1.03, P = 0.0373), 5WT < 9 (OR = 1.07, P = 0.0011) and CDT failure (OR = 1.04, P = 0.0088). A BMI < 21 kg/m^2^ was associated with a higher risk of pathologic MMSE (OR = 2.71, P = 0.0145) and verbal fluency (OR = 3.92, P = 0.0042) compared to a normal BMI (BMI 21–25).

**Table 3.** Factors associated with pathologic cognition and depression (multivariate analysis)

| Effect | Adjusted OR [95% CI] | Pr > Chi-Square |
| --- | --- | --- |
| **MMSE (N=681)** |  |  |
| Age | 1.03 [1.00; 1.06] | 0.0373 |
| Alcohol (yes vs. no) | 2.71 [1.22; 6.03] | 0.0145 |
| BMI < 21 vs. [21; 25] | 2.18 [1.25; 3.80] | 0.0063 |
| BMI > 30 vs. [21; 25] | 0.74 [0.49; 1.10] | 0.4914 |
| BMI ≥ 25; 30] vs. [21; 25] | 0.84 [0.51; 1.38] | 0.1345 |
| Educational level: intermediate vs. high | 1.77 [1.07; 2.93] | 0.0271 |
| **5WT (N=586)** |  |  |
| Age | 1.07 [1.03; 1.12] | 0.0011 |
| Educational level: low vs. intermediate or high | 2.21 [1.21; 4.02] | 0.0095 |
| Polypharmacy ≥ 5 (yes vs. no) | 0.52 [0.31; 0.89] | 0.0176 |
| Current smoker (yes vs. no) | 1.94 [1.07; 3.52] | 0.0299 |
| **Clock drawing test (N=624)** |  |  |
| Age | 1.04 [1.01; 1.07] | 0.0088 |
| Educational level: intermediate vs. high | 1.86 [1.11; 3.12] | 0.0188 |
| Educational level: low vs. intermediate or high | 2.07 [1.39; 3.09] | 0.0004 |
| Gender (males vs. females) | 0.68 [0.48; 0.96] | 0.0297 |
| **Fluency test (N=535)** |  |  |
| Hypertension (yes vs. no) | 1.67 [1.09; 2.57] | 0.0196 |
| BMI < 21 vs. [21; 25] | 3.92 [1.54; 9.99] | 0.0042 |
| BMI > 30 vs. [21; 25] | 1.04 [0.66; 1.65] | 0.3836 |
| BMI ≥ ]25; 30] vs. [21; 25] | 0.79 [0.46; 1.35] | 0.8624 |
| Educational level: intermediate vs. high | 1.91 [1.07; 3.41] | 0.0291 |
| Educational level: low vs. intermediate or high | 3.11 [1.99; 4.86] | <0.0001 |
| Gender (males vs. females) | 0.48 [0.32; 0.73] | 0.0005 |
| **GDS >9 (N=598)** |  |  |
| Polypharmacy ≥ 5 (yes vs. no) | 2.16 [1.40; 3.34] | 0.0005 |

The educational level (low *versus* intermediate or high educational level) was associated with a higher risk of 5WT < 9 (OR = 2.21, P = 0.0095), CDT failure (OR = 2.07, P = 0.0004), and pathologic verbal fluency (OR = 3.11, P < 0.0001). An intermediate educational level (*versus* high) was associated with a higher risk of pathologic MMSE (OR = 1.77, P = 0.0271), CDT failure (OR = 1.86, P = 0.0188), and pathologic verbal fluency (OR = 1.91, P = 0.0291).

Other variables independently associated with the risk of pathologic cognition were alcohol consumption (OR = 2.71, P = 0.0145) using the MMSE test, current smoking (OR = 1.94, P = 0.0299) using the 5WT test, and hypertension (OR = 1.67, P = 0.0196) using the fluency test. Conversely, males (*versus* females) had a lower risk of pathologic dysfunction using the CDT test (OR = 0.68, P = 0.0297) and the fluency test (OR = 0.48, P = 0.0005). Polypharmacy was associated with a lower risk of pathologic cognition using the 5WT (OR = 0.52, P = 0.0176). Polypharmacy was the only independent risk factor significantly associated with depression (GDS score > 9) (OR = 2.16; P = 0.0005).

## 4. Discussion

The preservation of cognitive functions is necessary to maintain community-dwelling older subjects in good health and to prevent or reduce loss of autonomy in this population. In our cohort of subjects ≥ 75 or ≥ 65 years with at least two comorbidities, but no diagnosed neurodegenerative disorder [Boyer, 2022], 34% had a pathologic global cognitive function when assessed using the MMSE adjusted for educational level, which was significantly higher (48%) in frail subjects compared to pre-frail subjects (30%) and robust subjects (15%). Moreover, 68% of subjects (77% of frail subjects) had verbal fluency impairment, principally literal fluency (60%) and less frequently categorial fluency (30%). This is suggestive of possible cognitive impairment [Faria, 2015]. In contrast, this population had no denomination problem on the 5WT, and only 17% (19% of frail subjects) had a 5WT score < 9. This score is indicative of suspicion of dementia or AD with a sensitivity of 80% and a specificity of 60% [Dubois, 2002]. The CDT is used to assess visuo-constructional ability and may be useful to detect pathologic cognition because it is less dependent on education or language than the MMSE test [Shulman, 2000]. The CDT is more performant than the MMSE to detect moderate to severe cognitive impairment in older community-dwelling individuals, and less performant to detect mild cognitive impairment [Nishiwaki, 2004]. Here, 46% of older subjects failed the CDT (59% of frail *versus* 26% of robust subjects). Cognitive performance is influenced by psychological factors, including anxiety and depressive mood, in older adults [Potvin, 2013; Shimada, 2014]. In our cohort, about half of the subjects had a GDS score > 9 (70% of frail *versus* 25% of robust) subjects) indicating depression in the previous week. Therefore, pathologic cognition was significantly more prevalent in frail older subjects, as reported previously [Robertson, 2014]. Nevertheless, even robust subjects may have cognitive impairment, especially when assessed by verbal fluency test.

Frailty and cognitive impairment are associated with functional decline. The ability to engage in ADL/IADL is dependent on cognitive ability, specifically executive functioning and memory [Lee, 2005; Suchy 2011; Connolly, 2017]. In our cohort, regardless of the cognitive test used, older subjects with low cognitive performance were significantly more frequently limited in ADL/IADL compared to subjects without cognitive dysfunction. This accounted for 64–69% of subjects with a cognitive deficit and limitation in ADL (*versus* 45–56% of subjects without cognitive deficit) and 83–94% of subjects with a limitation in IADL (*versus* 63–74%). In contrast, some older subjects without pathologic cognitive function may be also limited in ADL and IADL, likely due to impaired physical performance, sensory performance, or perception.

Cognitive function is associated with several interrelated confounding factors, including clinical and behavioral risk factors, health-related activity limitation, social isolation, and depressive symptoms. In this study, we assessed the associations of sociodemographic and clinical variables reportedly linked to cognitive impairment in community-dwelling older adults, including age, sex, BMI, education, cardiovascular risk factors (hypertension, dyslipidemia, and diabetes), polypharmacy, lifestyle factors (smoking and alcohol consumption), and social isolation/loneliness. Age was associated with loss of cognition in most psychometric tests (except the verbal fluency test), corresponding to brain aging and neurodegenerative decline [Murman, 2015]. Low educational level is also a risk factor for cognitive impairment, and a high educational level may be associated with increased cognitive reserve throughout life [Evans, 2019]. The risk of cognition impairment in our cohort was higher in subjects with a low BMI (< 21 kg/m^2^) in the MMSE and fluency test, which may be related to purported links of malnutrition, diet, and several nutrients with neurodegeneration and cognitive decline with age [Moore, 2018; Morley, 2014]. A large cross-sectional study involving more than 20,000 community-dwelling older adults (age ≥ 60 years) in Colombia revealed that underweight (BMI < 18.5 kg/m^2^) was associated with a decreased MMSE score and worse performance in the IADL. In contrast, being overweight was associated with increased MMSE and IADL scores [Borda, 2021]. Vasculature also contributes to cognitive impairment and dementia [Gorelik, 2011]. A recent Mayo Clinic study on aging confirmed the association of cardiovascular diseases and risk factors with cognitive decline after the age of 45–50 years, especially in females [Huo, 2022]. We found that hypertension was an independent covariate of poor performance on the fluency cognitive test. Although diabetes is reportedly a risk factor for frailty and cognitive impairment in subjects older than 70 years [Domergue, 2021], it was not an independent risk factor in our cohort.

Current or past or alcohol consumption is a secondary cause of cognitive impairment, and moderate drinking exerts detrimental effects on brain structure [Topiwala, 2017]. We found that alcohol consumption was independently associated with poor cognitive performance in the MMSE test. Chronic cigarette smoking is associated with adverse effects on vascular and brain function [Durazzo, 2007], and tobacco dependence is linked to cognitive impairment and dementia, as are other age-related disorders, including frailty [Durazzo, 2014; Kojima, 2015]. In the present study, current smoking was associated with pathologic cognition using the 5WT, which has a good positive predictive value for dementia [Dubois, 2002].

Although social isolation or loneliness among older adults may be a risk factor for cognitive impairment [Luchetti, 2020], living alone was not a significant risk factor for impaired cognitive function in the multivariate analysis. A recent longitudinal study in the United Kingdom revealed that living alone was not associated with poorer cognitive function at baseline or the 2-year follow-up in subjects ≥ 65 years of age; the authors speculated that such subjects might be more engaged in social activities [Evans, 2019]. Social activity may be a better covariate than living arrangement, but this was not fully recorded in this study. Polypharmacy was inversely associated with pathologic cognition using the 5WT. This result is controversial and seems to be due to the low sensitivity of the 5WT to detect a dementia. In our study, a significant number of polypharmacy subjects were not detected by the 5WT. So what we have to remember is that the link between polypharmacy and cognitive impairment is not clear, and probably more the use of certain categories of drugs such as anticholinergics, benzodiazepines, which explains the link with the classical memory tests (except 5WT) [Pieper, 2020 & Eshetie 2019 & Penninkilampi 2018].

This study had several limitations. Due to its cross-sectional design, the causality of independent covariates of poor cognitive performance could not be established. Longitudinal studies will be necessary to confirm the associations of cofactors with pathologic cognitive impairment. Additionally, cognitive performance measured by psychometric tests may not be specific in subjects with depression. In another cross-sectional analysis of the FREEDOM-LNA cohort, age, depression, impaired cognition, and diabetes were significantly associated with frailty [Boyer, 2022]. Also, poor cognitive performance using the MMSE was associated with the trajectory of frailty, independently of depression [Boyer, 2022].

Another limitation is the missing data for some subjects, which could affect the differences between the MMSE and the other psychometric tests. Furthermore, the cohort was composed of older subjects who asked to be visited at home for a comprehensive geriatric assessment [Boyer, 2022]. Therefore, most of the subjects were frail or pre-frail and had several comorbidities, and thus may not be representative of the general population of community-dwelling older subjects.

We identified several potentially modifiable risk factors associated with pathologic cognitive performance in community-dwelling older subjects. Maintaining a normal BMI, managing hypertension, and stopping alcohol consumption and smoking may be included in interventions to prevent or improve cognitive decline, frailty, and disability. In line with the WHO recommendation [WHO, 2019], interventions to promote cessation of smoking and/or alcohol consumption may reduce the risk of cognitive decline and dementia. Additionally, physical, nutritional, and cognitive interventional approaches can reverse frailty among community-living older subjects [Ng, 2015]. A randomized study involving subjects of mean age 75 years revealed that brain exercise could improve cognitive function [Htut, 2018]. A recent systematic review confirmed that combined cognitive and physical exercise training was more advantageous than cognitive training in terms of attention, memory, language, processing speed, and global cognition [Rieker, 2022].

In conclusion, the prevalence of cognitive impairment among frail subjects was high, and older subjects were more frequently affected in ADL. Among this study population, a low BMI, hypertension, alcohol consumption, and smoking were significantly associated with pathologic cognitive dysfunction after adjustment for age, educational level, living alone, and other cardiovascular risk factors.

## Statements

## Data Availability

The data underlying the results presented in this study contain potentially identifiable personal information and cannot be made publicly available due to ethical and legal restrictions, including compliance with French data protection regulations (CNIL) and the conditions of participant consent. Researchers who meet the criteria for access to confidential data may request access to the FREEDOM cohort data from the FREEDOM study steering committee by contacting Dr Caroline Gayot (). Requests will be evaluated in accordance with ethical approvals, data protection regulations, and institutional policies.

## Acknowledgement

The authors would like to thank study participants, Axonal-Biostatem (Castries, France) for data management and statistics, and Thierry Radeau Consulting (Epinay-Sous-Senart, France), the University of Limoges Partenariale Foundation and “Carsat Centre Ouest” de Limoges

## Author Contributions

LM, KR, and AT drafted the manuscript. AT, AL, BF, ND, KR, MDC and LM read and revised the manuscript. AT, KR, AL and BF helped perform statistical analysis. AT, NC, CG and CLM collected data. AT, CG, MDC and KR participated in the design of the study methodology and helped draft the manuscript. All authors read and approved the final manuscript.

## Availability of data and materials

Doctor Caroline Gayot, PhD who should be contacted if someone wants to request the data. Data are not publicly available due to privacy or ethical restrictions.

## Abbreviations

BMI: body mass index
MMSE: Mini Mental State Examination
ADL: activities of daily living
IADL: instrumental activities of daily living
5WT: 5-word test
GDS: geriatric depression scale
CDT: clock drawing test
OR: odds ratio

